# Double burden of malnutrition and cardiometabolic risk among older residents of a state social-care institution in Kazakhstan: a cross-sectional study

**DOI:** 10.64898/2026.07.09.26357706

**Authors:** Aigul Abduldayeva, Saule Iskakova, Gulnur Doszhanova, Olzhas Kozhamkulov, Saule Tardzhibayeva, Zhanar Bukeyeva, Aida Shuakbayeva, Karlygash Suindik, Yerkezhan Tulegenova, Zhamal Lenzatova, Anara Aktanova

## Abstract

Older adults in residential care are usually described as a group at high risk of undernutrition, yet data on the nutritional and cardiometabolic status of institutionalised older adults in Central Asia are scarce. We aimed to characterise the anthropometric, dietary and biochemical profile of older residents of a state social-care institution in Kazakhstan and to examine whether overnutrition, micronutrient inadequacy and cardiometabolic risk coexist. In this cross-sectional study, 62 adults aged 60 years and over from the “Sharapat” centre in Astana underwent anthropometry, bioelectrical impedance body-composition analysis, blood-pressure measurement, dietary assessment (specialized nutrition questionnaires, a 24-hour dietary recall and food diaries) with calculation of nutrient intakes, and venous blood and urine testing; serum 25-hydroxyvitamin D and trace elements were measured in 29 participants. Laboratory analyses and data processing were performed at the Research Institute of Preventive Medicine named after E.D. Dalenov, Astana Medical University. The sample (61.3% men; mean age 74.0 years) showed a high cardiometabolic burden, with arterial hypertension in 63%, total cholesterol of at least 5.0 mmol/L in 60%, LDL-cholesterol of at least 3.0 mmol/L in 71%, and overweight or obesity in 58%, whereas only 5% were underweight. Habitual diets were high in sodium (71% above 2000 mg/day) and low in potassium (92% below 3500 mg/day), calcium (85% below 1000 mg/day) and fibre (90% below 25 g/day). Among those tested, 79% had vitamin D deficiency, and overweight or obesity coexisted with vitamin D deficiency in 16 of 29 participants. None of 56 exploratory diet-risk correlations survived correction for multiple testing. Rather than the undernutrition typical of residential care, these residents displayed a double burden of malnutrition-excess adiposity and cardiometabolic risk alongside micronutrient-poor diets and widespread vitamin D deficiency-identifying concrete targets for institutional catering, supplementation and cardiometabolic screening.

## Introduction

Populations are ageing rapidly worldwide, and the nutritional status of older adults has become a central determinant of healthy ageing and of the burden of non-communicable disease. In residential and long-term care, the problem most often emphasised in the international literature is undernutrition: a systematic review and meta-analysis using the Mini Nutritional Assessment found that malnutrition and its risk increase with the level of care and reach their highest values in nursing homes and long-term care, far exceeding the values seen in community-dwelling older people [1]. Frailty, weight loss, dysphagia and feeding dependence dominate this narrative.

A different problem has emerged in parallel. The double burden of malnutrition-the coexistence of overnutrition with undernutrition and micronutrient deficiency—is increasingly recognised not only across populations but within individuals, including older adults with obesity, in whom excess adiposity may mask sarcopenia and specific nutrient deficiencies [2]. Whether institutionalised older adults in transitional, middle-income settings follow the classic undernutrition pattern or this newer double-burden pattern is largely unknown.

Kazakhstan, like other Central Asian countries, carries a high and rising burden of cardiometabolic disease. Population studies in the Astana region have reported arterial hypertension in approximately 70% of adults aged 50–75 years [3], together with a high prevalence of dyslipidaemia [4] and diabetes [5] in the same population, and national data confirm widespread hypercholesterolaemia [6]. Vitamin D deficiency is also common, with a pooled prevalence of approximately 57% among Kazakh adults [7]. Despite this, the nutritional and cardiometabolic status of older adults living in state social-care institutions in the region has not been described.

Using an integrated assessment of anthropometry, body composition, habitual diet and biochemistry in residents of a state social-care centre in Astana, we aimed to characterise their cardiometabolic and nutritional profile and to test whether overnutrition, micronutrient inadequacy and cardiometabolic risk coexist in this setting.

## Materials and methods

### Study design and setting

This was a cross-sectional, observational study conducted among older adults residing at the “Sharapat” Centre for Social Services of the Akimat of Astana (Akkorgan St. 2, Astana), a state medico-social institution for older people. Anthropometric data and questionnaire responses were collected on site; laboratory analyses and statistical data processing were conducted at the academician E.D. Dalenov Research Institute of Preventive Medicine at the Astana Medical University.

The study is reported in accordance with the STROBE guidelines for observational studies (S1 Checklist).

### Ethics statement

The study was conducted in accordance with the Declaration of Helsinki and was approved by the Local Bioethics Committee of NJSC “Astana Medical University” (decision No. 15/ID:169 of 28 April 2026; valid until 28 April 2027). All participants provided written informed consent before enrolment. As older adults receiving institutional care are considered a potentially vulnerable population, additional safeguards were applied: consent was obtained directly from each participant without involvement of institutional staff in the consent process, participants could withdraw at any time without any effect on the care or services they received at the institution, and participation had no bearing on the care or services the participant received at the institution.

#### Participants

Recruitment was conducted using a continuous (accessible) sampling method of residents who met the selection criteria during the data collection period (05.January–26.June 2026). Inclusion criteria: age 60 years or older, residence in a medical-social center, and signed written informed consent. Exclusion criteria: age under 60 years, history of diagnosed cancer, allergic, or autoimmune diseases, incomplete questionnaire, or lack of signed informed consent. Initially, 69 residents were screened; 7 were excluded due to incomplete questionnaires, resulting in 62 participants who met all criteria being included in the analysis.

### Anthropometry and body composition

Height, body weight, and waist and hip circumference were measured using standard procedures, and body mass index (BMI) and the waist-to-hip ratio were calculated. Body composition-total and visceral fat, skeletal muscle mass, basal metabolic rate, body water and bone mass-was assessed by bioelectrical impedance analysis. An appendicular lean mass index (ALMI) was derived as skeletal muscle mass divided by height squared; sarcopenia was screened using the revised European Working Group on Sarcopenia in Older People (EWGSOP2) ALMI cut-offs of below 7.0 kg/m^2^ in men and below 5.5 kg/m^2^ in women [8], with the caveat noted in the Limitations.

### Blood pressure

Systolic and diastolic blood pressure and heart rate were recorded under standardised conditions. Arterial hypertension was defined as systolic blood pressure of at least 140 mmHg and/or diastolic blood pressure of at least 90 mmHg.

### Dietary assessment

Habitual diet and nutritional status were assessed using specialized nutrition questionnaires, a 24-hour dietary recall and food diaries covering culturally specific foods (including horse meat, kurt, kymyz, bauyrsak and traditional broths), from which daily intakes of energy, macronutrients, sugar, fibre and selected minerals and vitamins were computed. Reference thresholds applied were sodium above 2000 mg/day, potassium below 3500 mg/day, calcium below 1000 mg/day and fibre below 25 g/day; the dietary sodium-to-potassium mass ratio was calculated, given its reported relationship with cardiometabolic risk and cardiovascular outcomes [9,10].

### Laboratory analyses

Venous blood and urine samples were analysed for a complete blood count, biochemistry (total protein, alanine and aspartate aminotransferases, gamma-glutamyl transferase, alkaline phosphatase, total and direct bilirubin, urea, creatinine and uric acid), a fasting lipid profile (total, LDL- and HDL-cholesterol) and fasting venous glucose, and urinalysis. Serum 25-hydroxyvitamin D and trace elements (including selenium and zinc) were measured in a subset of 29 participants. Hypercholesterolaemia was defined as total cholesterol of at least 5.0 mmol/L, elevated LDL-cholesterol as at least 3.0 mmol/L, low HDL-cholesterol as below 1.0 mmol/L in men and below 1.3 mmol/L in women, and hyperglycaemia as fasting glucose of at least 6.1 mmol/L. Vitamin D status was classified as deficient (below 20 ng/mL) or insufficient (below 30 ng/mL).

### Statistical analysis

Statistical analysis was performed in IBM SPSS Statistics 31.0 (IBM Corp., Armonk, NY, USA). Continuous variables are presented as mean ± SD or median [Q1; Q3] and categorical variables as counts and percentages, overall and by sex; between-sex differences were tested with the Mann– Whitney U test. Associations between diet and cardiometabolic markers were explored with Spearman correlations using a Bonferroni-corrected threshold and were treated as hypothesis-generating. Two-sided p-values below 0.05 were considered nominally significant.

## Results

### Characteristics of the study population

Of the 62 participants, 38 (61.3%) were men and 24 (38.7%) were women; the median age was 73.5 years (IQR 68.2–78.0; range 60–90). By the World Health Organization classification, 33 (53.2%) were in the elderly group (60–74 years) and 29 (46.8%) in the senile group (75–90 years); women were older than men. The sample was male-predominant, which is atypical for residential-care populations. Demographic, anthropometric, body-composition and cardiometabolic characteristics are shown in Table 1.

**Table 1.** Demographic, anthropometric, body-composition and cardiometabolic characteristics, overall and by sex (n = 62).

| Variable | Unit | n | Overall, median (IQR) | Men | Women | p* |
| --- | --- | --- | --- | --- | --- | --- |
| Age | years | 62 | 73.5 (68.2–78.0) | 71.5 (67.2–75.8) | 78.0 (73.0–84.0) | 0.001 |
| Height | cm | 62 | 163.5 (154.0–171.0) | 168.5 (163.2–173.0) | 152.0 (149.5–156.2) | <0.001 |
| Weight | kg | 62 | 69.3 (62.2–78.1) | 71.1 (65.6–77.4) | 66.2 (61.0–80.2) | 0.363 |
| Body mass index | kg/m<sup>2</sup> | 62 | 26.1 (23.1–28.9) | 24.2 (22.1–26.8) | 28.4 (26.0–32.6) | 0.003 |
| Waist circumference | cm | 62 | 95.0 (89.2–105.6) | 94.2 (88.0–104.1) | 95.5 (91.8–107.2) | 0.312 |
| Hip circumference | cm | 62 | 103.0 (97.2–109.8) | 101.0 (94.2–107.1) | 104.0 (101.5–116.0) | 0.023 |
| Waist-to-hip ratio |  | 62 | 0.9 (0.9–1.0) | 1.0 (0.9–1.0) | 0.9 (0.9–0.9) | 0.018 |
| Systolic blood pressure | mmHg | 62 | 142.0 (127.5–164.5) | 135.0 (126.0–148.8) | 158.0 (133.0–171.0) | 0.021 |
| Diastolic blood pressure | mmHg | 62 | 89.0 (76.2–96.0) | 83.0 (73.5–96.0) | 91.5 (79.8–100.0) | 0.122 |
| Heart rate | bpm | 62 | 73.0 (67.0–83.0) | 73.0 (65.5–82.5) | 73.5 (69.2–83.2) | 0.659 |
| Total body fat | % | 62 | 33.0 (27.3–38.7) | 28.8 (24.2–33.8) | 39.1 (34.1–44.3) | <0.001 |
| Visceral fat | units | 62 | 14.5 (10.6–16.5) | 15.2 (11.6–18.5) | 11.2 (9.5–15.0) | 0.003 |
| Skeletal muscle mass | kg | 61 | 44.1 (38.4–49.9) | 47.6 (43.7–51.1) | 39.1 (35.4–42.4) | <0.001 |
| ALMI | kg/m<sup>2</sup> | 61 | 16.9 (15.4–18.1) | 16.9 (15.7–18.2) | 16.9 (15.2–17.8) | 0.842 |
| Body water | % | 62 | 47.5 (43.8–51.4) | 49.8 (45.8–53.1) | 43.4 (39.8–46.7) | <0.001 |
| Total cholesterol | mmol/L | 55 | 5.2 (4.6–5.8) | 5.1 (4.4–5.7) | 5.3 (5.0–6.1) | 0.151 |
| LDL-cholesterol | mmol/L | 55 | 3.6 (2.9–4.1) | 3.5 (2.7–4.1) | 3.7 (3.1–4.4) | 0.255 |
| HDL-cholesterol | mmol/L | 55 | 1.3 (1.1–1.5) | 1.3 (1.1–1.5) | 1.3 (1.1–1.6) | 0.813 |
| Fasting glucose | mmol/L | 55 | 5.0 (4.6–5.9) | 5.0 (4.6–5.9) | 5.0 (4.7–6.0) | 0.958 |
| Uric acid | μmol/L | 55 | 337.0 (273.8–382.8) | 347.2 (282.3–389.2) | 319.4 (240.1–358.8) | 0.164 |
| Haemoglobin | g/L | 55 | 140.0 (127.0–150.0) | 144.0 (135.5–151.0) | 133.0 (120.2–140.0) | 0.012 |

p\*, Mann–Whitney U test (men vs women); IQR, interquartile range; ALMI, appendicular lean mass index.

Women had a significantly higher BMI than men (median 28.4 vs 24.2 kg/m^2^; p = 0.003) and higher total body fat, consistent with the sex difference in adiposity.

### Cardiometabolic burden

The prevalence of cardiometabolic conditions is summarised in Table 2. Overweight or obesity affected 58% of participants and abdominal obesity 55%, whereas only 5% were underweight. Arterial hypertension was present in 63%, hypercholesterolaemia in 60%, elevated LDL-cholesterol in 71%, low HDL-cholesterol in 31%, hyperglycaemia in 24% and anaemia in 22%.

**Table 2.**
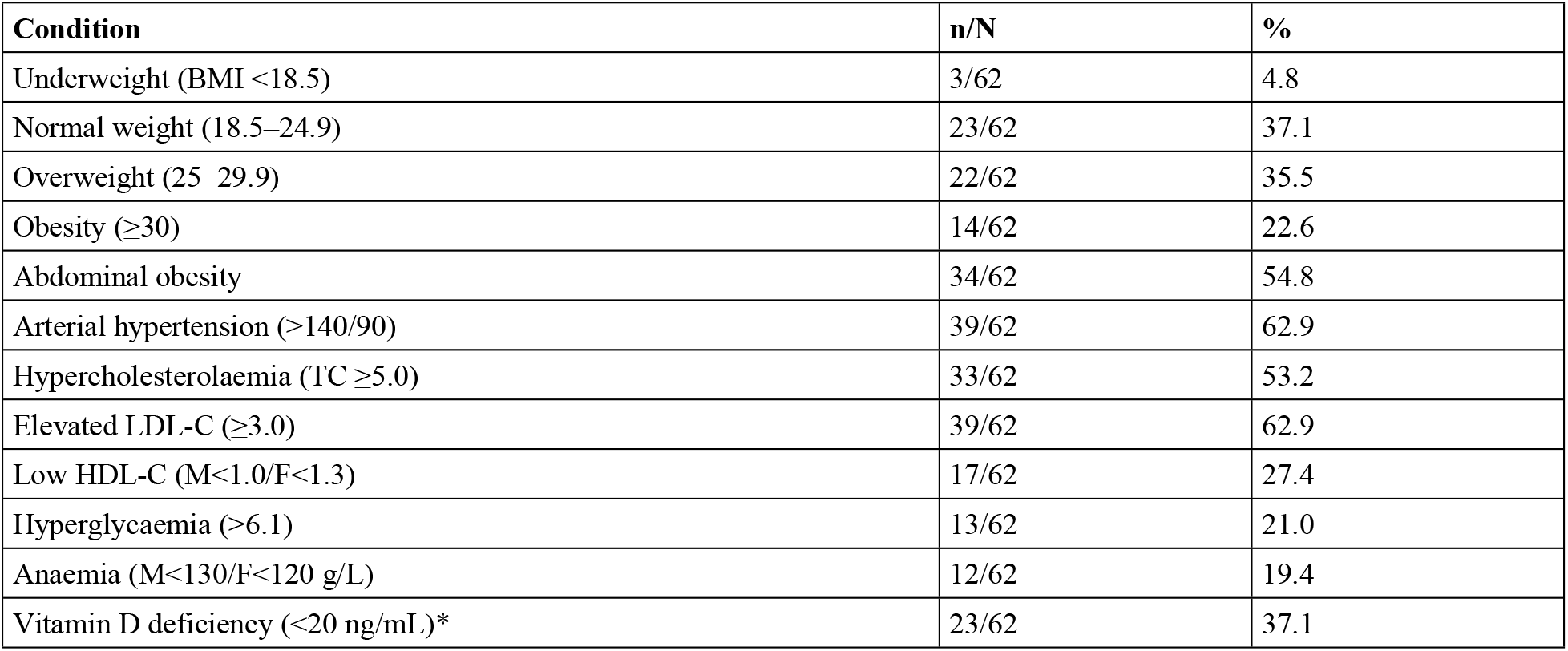
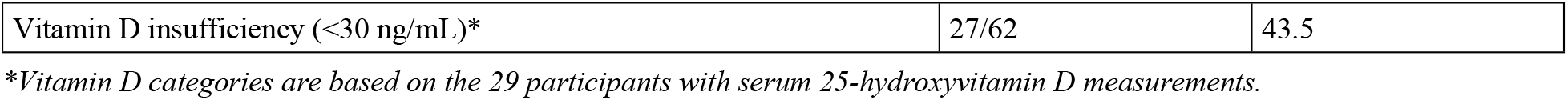
Prevalence of cardiometabolic and nutritional conditions (n = 62).

| Condition | n/N | % |
| --- | --- | --- |
| Underweight (BMI <18.5) | 3/62 | 4.8 |
| Normal weight (18.5–24.9) | 23/62 | 37.1 |
| Overweight (25–29.9) | 22/62 | 35.5 |
| Obesity (≥30) | 14/62 | 22.6 |
| Abdominal obesity | 34/62 | 54.8 |
| Arterial hypertension (≥140/90) | 39/62 | 62.9 |
| Hypercholesterolaemia (TC ≥5.0) | 33/62 | 53.2 |
| Elevated LDL-C (≥3.0) | 39/62 | 62.9 |
| Low HDL-C (M<1.0/F<1.3) | 17/62 | 27.4 |
| Hyperglycaemia (≥6.1) | 13/62 | 21.0 |
| Anaemia (M<130/F<120 g/L) | 12/62 | 19.4 |
| Vitamin D deficiency (<20 ng/mL)* | 23/62 | 37.1 |
| Vitamin D insufficiency (<30 ng/mL)* | 27/62 | 43.5 |

\*Vitamin D categories are based on the 29 participants with serum 25-hydroxyvitamin D measurements.

### Dietary intake

Dietary intakes and the proportion of participants outside reference values are shown in Table 3. Diets were high in sodium (median 2452 mg/day; 71% above 2000 mg/day) and low in potassium (median 2095 mg/day; 92% below 3500 mg/day), giving a median sodium-to-potassium ratio of 1.26. Calcium intake was below 1000 mg/day in 85% of participants and fibre intake below 25 g/day in 90%.

**Table 3.** Daily dietary intake and proportion of participants outside reference values (n = 62).

| Nutrient | Unit | Median (IQR) | Outside reference |
| --- | --- | --- | --- |
| Energy | kcal/day | 1574.0 (1308.3–1970.2) | – |
| Protein | g/day | 62.3 (51.8–73.9) | – |
| Total fat | g/day | 63.2 (53.9–72.1) | – |
| Saturated fat | g/day | 24.3 (20.2–30.9) | – |
| Carbohydrate | g/day | 201.4 (148.4–256.2) | – |
| Sugar | g/day | 49.1 (32.0–75.4) | – |
| Dietary fibre | g/day | 12.7 (9.3–18.7) | 90% (<25) |
| Sodium | mg/day | 2451.6 (1939.5–3204.0) | 71% (>2000) |
| Potassium | mg/day | 2094.7 (1655.4–2510.2) | 92% (<3500) |
| Sodium-to-potassium ratio |  | 1.26 (0.85–1.49) | – |
| Calcium | mg/day | 563.0 (352.1–833.7) | 85% (<1000) |
| Magnesium | mg/day | 246.5 (180.8–314.8) | – |
| Iron | mg/day | 12.1 (8.9–15.7) | – |

Reference thresholds: fibre <25 g/day; sodium >2000 mg/day; potassium <3500 mg/day; calcium <1000 mg/day.

### Vitamin D and micronutrient status

Among the 29 participants with serum measurements, the median 25-hydroxyvitamin D concentration was 11.7 ng/mL; vitamin D deficiency (below 20 ng/mL) was present in 23 of 29 (79%) and insufficiency (below 30 ng/mL) in 27 of 29 (93%). Overweight or obesity coexisted with vitamin D deficiency in 16 of these 29 participants.

### Exploratory diet–risk associations

Of 56 pairwise Spearman correlations between eight dietary variables and seven cardiometabolic markers, only the association between potassium intake and fasting glucose was nominally significant (rho = 0.32; p = 0.017; n = 55) and did not survive Bonferroni correction. No other correlation reached significance, consistent with limited statistical power rather than a definitive absence of association.

## Discussion

In this group of older adults in a state social-care institution in Kazakhstan, the nutritional picture departed markedly from the undernutrition that dominates the international residential-care literature, in which malnutrition rises with the level of care and is highest in nursing homes [1]. Here, underweight was rare (5%), whereas overweight and obesity were common (58%) and were accompanied by a high prevalence of hypertension, hypercholesterolaemia and elevated LDL-cholesterol. At the same time, habitual diets were poor in potassium, calcium and fibre and high in sodium, and vitamin D deficiency was almost universal among those tested. This constellation-excess adiposity and cardiometabolic risk coexisting with micronutrient inadequacy-is best understood as a double burden of malnutrition, a concept increasingly applied at the individual level and to older adults with obesity [2].

The cardiometabolic findings are consistent with previous regional data. Hypertension affected 63% of our participants, in keeping with the high prevalence reported among older adults in the Astana region [3], and the high frequency of elevated cholesterol mirrors regional and national reports of dyslipidaemia and hypercholesterolaemia [4,6]. Our results extend these observations to a previously undescribed group-older residents of state social care-and indicate that institutional placement does not protect against, and may coincide with, a substantial cardiometabolic burden.

The dietary pattern is itself informative. The combination of high sodium and low potassium, captured by the sodium-to-potassium ratio, is relevant because this ratio has been associated with hypertension and metabolic syndrome in older Korean adults [9] and with stroke, cardiovascular disease and mortality in a Japanese cohort [10]. Low potassium, calcium and fibre intakes point to a diet limited in fruit, vegetables and dairy and identify specific, modifiable targets. Because, in this setting, food is largely provided by the institution, these inadequacies are directly actionable through catering standards.

The very high prevalence of vitamin D deficiency (79% among those tested) exceeded the pooled adult estimate of approximately 57% for Kazakhstan [7], consistent with the expectation that older, institutionalised individuals with limited sun exposure are at especially high risk. Together with the low dietary calcium and the observed anaemia, this supports routine assessment and supplementation protocols within such facilities.

The male predominance of the sample (61%) is atypical for residential care, where women usually predominate, and may reflect the specific pathways by which older adults enter state social care in Kazakhstan. This demographic feature warrants attention in its own right and when interpreting sex-specific findings, such as the higher BMI observed in women.

We did not detect robust direct associations between individual dietary components and cardiometabolic markers. This most plausibly reflects the limited power of a small sample, the measurement error inherent in food-frequency questionnaires, and the cross-sectional design, rather than a true absence of relationships; the results should therefore be read as a descriptive characterisation that motivates larger, adequately powered studies.

### Strengths and limitations

The principal strength of this study is the integration, for a single previously undescribed group, of anthropometry, bioimpedance body composition, a culturally tailored dietary assessment and a broad biochemical panel. Limitations include the small, single-institution sample (n = 62), which limits power and generalisability; the cross-sectional design, which precludes causal inference; reliance on self-reported dietary assessment, with wide intake ranges suggesting some reporting error at the extremes; the availability of serum micronutrient and vitamin D data for only 47% of participants, making those estimates exploratory; and calculation of the ALMI from total rather than appendicular lean mass, so that the sarcopenia classification, which relies on appendicular cut-offs [8], should be confirmed with validated methods.

## Conclusions

Older residents of a state social-care institution in Kazakhstan showed a double burden of malnutrition: a high prevalence of hypertension, dyslipidaemia and excess adiposity together with sodium-rich, micronutrient-poor diets and widespread vitamin D deficiency, rather than the undernutrition typically described in residential care. These findings define concrete targets for institutional catering, vitamin D supplementation and cardiometabolic screening, and underline the need for larger, prospective studies of institutionalised older adults in Central Asia.

## Data Availability

Data cannot be shared publicly because they contain potentially identifying and sensitive information on a vulnerable population (institutionalised older adults), and public deposition was not covered by participants' informed consent or by the approval of the Local Bioethics Committee of NJSC "Astana Medical University". De-identified participant-level data are available upon reasonable request, for researchers who meet the criteria for access to confidential data, from the Local Bioethics Committee of NJSC "Astana Medical University" (Contact: [ / +7753867942—phone number of the Ethics Committee secretary]).

## Acknowledgments

The authors thank the residents and staff of the “Sharapat” Centre for the Provision of Special Social Services for their participation and support.

## Author contributions

Conceptualization: Saule Iskakova. Project administration: Aigul Abduldayeva. Supervision: Zhanar Bukeyeva. Investigation (data and material collection): Olzhas Kozhamkulov, Aida Shuakbayeva, Karlygash Suindik, Yerkezhan Tulegenova, Zhamal Lenzatova, Anara Aktanova.

Data curation (database assembly): Olzhas Kozhamkulov. Formal analysis: Gulnur Doszhanova, Saule Tardzhibayeva. Writing – original draft: Gulnur Doszhanova, Saule Tardzhibayeva.

Writing – review & editing: Saule Iskakova (final version); all authors.

All authors have read and approved the final version of the manuscript.

## Supporting information

**S1 Checklist**. STROBE checklist for cross-sectional studies.

**S1 Dataset**. De-identified participant-level dataset (anthropometry, body composition, dietary intakes and laboratory results).

**S1 File**. Data dictionary describing variables, units and coding for the dataset.

## Notes

### Competing Interest Statement

The authors have declared no competing interest.

### Author Declarations

The study was approved by the Local Bioethics Committee of NJSC “Astana Medical University” (decision No. 15/ID:169 of 28 April 2026). Written informed consent was obtained from all participants. The study conformed to the Declaration of Helsinki

